# Lyme disease increases risk for multiple gynecological conditions

**DOI:** 10.1101/2025.03.03.25323258

**Authors:** Paige S. Hansen Colburn, Grace Blacker, Sarah Galloway, Qingying Feng, Prasanna S. Padmanabham, Guido Pisani, Brandon T. Lee, Grace Loeser, Monika W Perez, Kunzan Liu, Jade Kuan, Emelia von Saltza, Satu Strausz, Lisa M. Mattei, Sophie VanderWeele, George R. Nahass, Amie Kitjasateanphun, Rangarajan Bharadwaj, Hari-Hara SK Potula, Maia Atzmon Shoham, Finngen, Victoria L. Mascetti, Eric Gars, Hanna M Ollila, Kaylon L. Bruner-Tran, Irving Weissman, Sixian You, Beth Pollack, Linda Griffith, Nasa Sinnott-Armstrong, Michal Caspi Tal

**Author notes:** Corresponding authors: Michal Caspi Tal and Nasa Sinnott-Armstrong.

## Abstract

Lyme disease (LD) is an illness caused by the spirochete *Borrelia burgdorferi* (*B. burgdorferi*). *Borrelia* is known to disseminate through organs, including the skin, joints, spinal cord, bladder, and heart, leading to Lyme arthritis, neuroborreliosis, and Lyme carditis. While previous studies have investigated the impact of LD on pregnancy in both mice and humans and have found the presence of *B. burgdorferi* in the uterus of mice, we studied the impact of LD on the non-pregnant female reproductive tract. We use a mouse model for LD and find an ongoing and severe infection of the reproductive tract of female mice, which persists up to 15-months post-inoculation. This infection results in uterine glandular cysts and endometrial hyperplasia as well as vaginal epithelial thickening, polymorphonuclear and mononuclear cell epithelial infiltration, and epithelial desquamation into the vaginal lumen. Strikingly, we find that age has an impact on the extent of gynecologic pathology such that aged female mice (1-year old) that are reproductively senescent have more gynecologic pathology with infection compared to young mice (15-weeks old) when infected for the same length of time. Using large-scale electronic healthcare record data, we report that LD additionally results in increased infection-associated risk of menorrhagia (1.5-fold), miscarriage (1.62-fold), uterine fibroids (1.42-fold), and endometriosis (1.93-fold). Underreporting of gynecological outcomes is pervasive throughout many different infectious diseases, and LD-associated gynecological pathologies may have been similarly underappreciated in the field. This work suggests that further study of the female reproductive tract and the effects of *B. burgdorferi* infection therein will help clarify and expand the knowledge of myriad LD outcomes.

**Graphical abstract:** 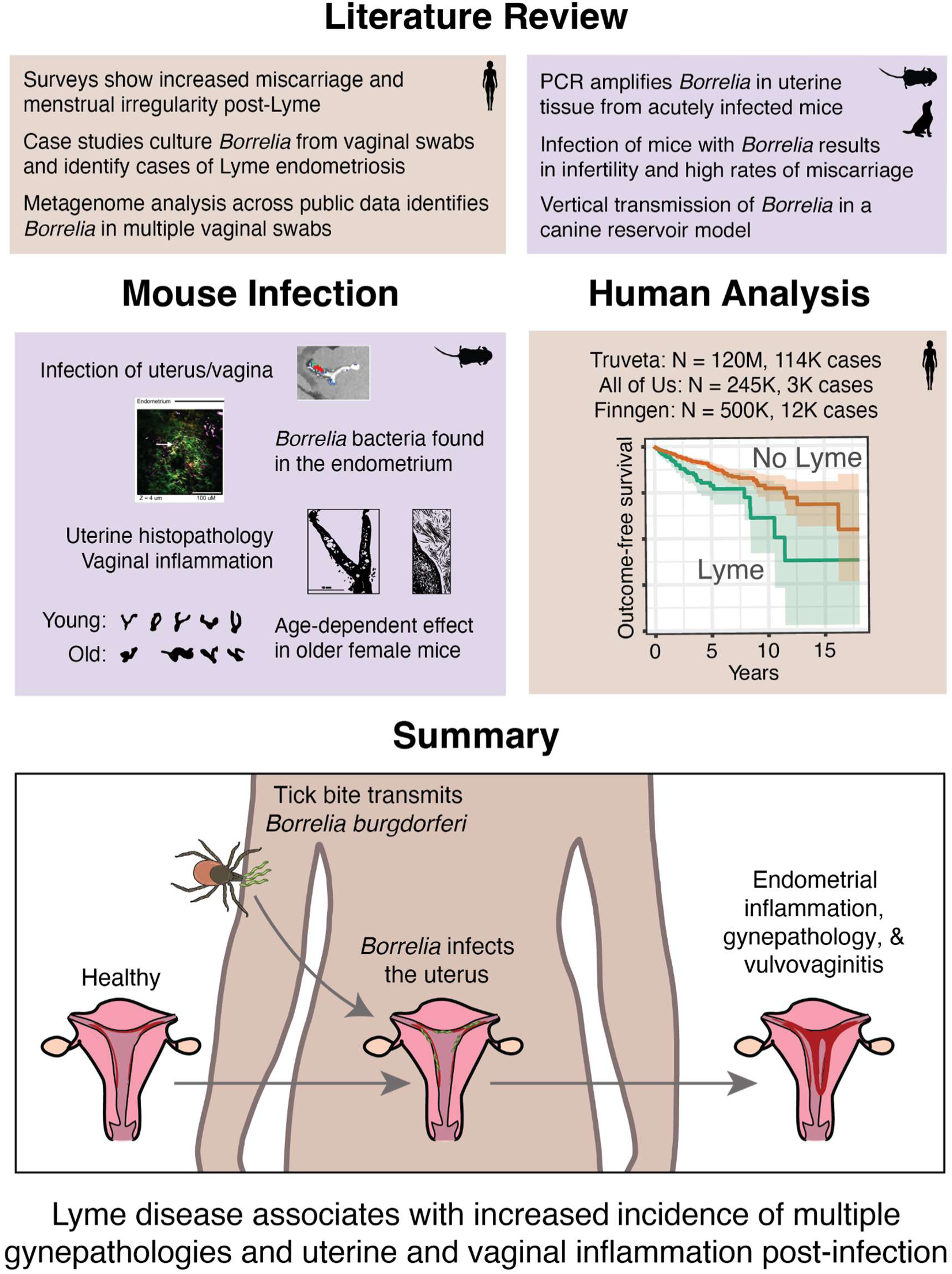

## Main

Infection with the spirochete *Borrelia burgdorferi* (*B. burgdorferi*), can lead to Lyme disease (LD)^1^. In addition to acute erythema migrans (EM) rash and flu-like symptoms, *B. burgdorferi* has been shown to infect the skin, joints, bladder, and heart^2–4^. These conditions may progress to a number of secondary outcomes, including Lyme arthritis, neuroborreliosis, and Lyme carditis^5^. However, the systemic nature of *B. burgdorferi* infection suggests that additional tissues might be susceptible to disease. For example, bacteria that are found in the bladder are also often found in the female reproductive tract ^6–10^, potentially contributing to the development of gynecologic disorders.

A different *Borrelia* species, *Borrelia crocidurae*, that causes tick borne relapsing fever has been implicated in complications with pregnancy^11,12^. Another pathogenic spirochetal bacteria *Treponema pallidum,* the bacteria that causes syphilis, is morphologically and phylogenetically similar to *B. burgdorferi,* colonizes the reproductive tract, and if left untreated causes reproductive organ damage such as syphilitic endometritis and miscarriage^13–16^. Building upon early work on increased rates of adverse birth outcomes from mothers with LD^17^, case studies of endometriosis and infertility associated with LD^18^, as well as the acute effect of *B. burgdorferi* in pregnancy through uterine acute infection^19^, we hypothesized that *B. burgdorferi* might increase risk for gynecological disease. Previous research has shown that infecting LD-symptomatic C3H mice 4-days after mating leads to increased fetal death, whereas infection 3-weeks prior to pregnancy did not. PCR analysis detected *B. burgdorferi* DNA in the uteri of the 4-days after mating infected mice, but not in the 3-weeks pre-mating infected mice (n=2) and rarely in the fetal tissues^19,20^. Vertical transmission of *B. burgdorferi* has been reported in dogs, where 8 out of 10 female beagles inoculated with *B. burgdorferi* prior to breeding, had at least 1 pup from each litter with evidence of *B. burgdorferi*, and pups from 2 litters yielded viable *B. burgorferi* by culture^21^. In addition, we examined data from previous studies of LD patients^22,23^, who reported strikingly elevated rates of miscarriage (Extended Data Figure 1A) and menstrual irregularity (Extended Data Figure 1B).

**Extended Figure 1.**
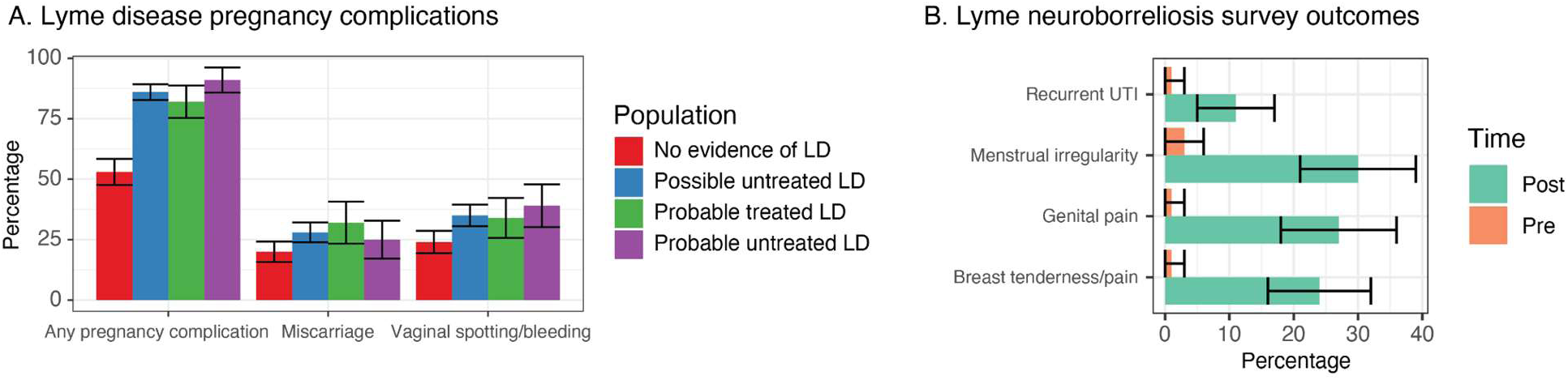
Existing evidence of obstetric and gynecological effects of Lyme disease (LD). A. Individuals who were pregnant with various experiences with LD reported adverse pregnancy outcomes and symptoms^22^. B. Individuals who developed Lyme neuroborreliosis reported increased rates of menstrual irregularity, recurrent UTIs, genital pain, and breast tenderness post infection^23^.

Together, these studies on reproductive outcomes also suggest a potential connection between gynecological pathologies and *B. burgdorferi* infection, and prompted us to further evaluate gynecological outcomes in both mice and humans.

### *B. burgdorferi* is present in the reproductive tract of infected mice

To examine why there is linkage between LD and other uterine disorders, irrespective of pregnancy, we used a murine infection model. To investigate if *B. burgdorferi* infects the reproductive tract of female mice post-*B. burgdorferi* infection, we infected female C57BL/6 (B6) mice intradermally with a N40-Luciferase strain of *B. burgdorferi* (luc-*B. burgdorferi*) and left them untreated for 8 weeks. *In Vivo* Imaging System (IVIS) images of the excised uteruses from infected mice were found to be luciferase-positive through bioluminescence (Figure 1A). The signal location in the female reproductive tract that luc-*B. burgdorferi* emanated from varied from animal to animal and across experiments, with bioluminescence noted in both the uterus and ovaries. Additionally, luc-*B. burgdorferi* are visualized via multiphoton imaging of the uterus with the highest luciferase signal from Figure 1A (Figure 1B). Upon long-term Green Fluorescence Protein-*B. burgdorferi* (GFP-*B. burgdorferi*) infection of B6 mice, digested reproductive tracts (including uterus, cervix, ovaries, and fallopian tubes) of infected mice were found to have GFP^+^ cells via flow cytometry whereas uninfected mice did not (Figure 1C). GFP^+^ sorted cells from reproductive tract digestions of infected B6 mice were found to have intracellular GFP, which was visualized via fluorescent confocal microscopy (Figure 1D).

**Figure 1.**
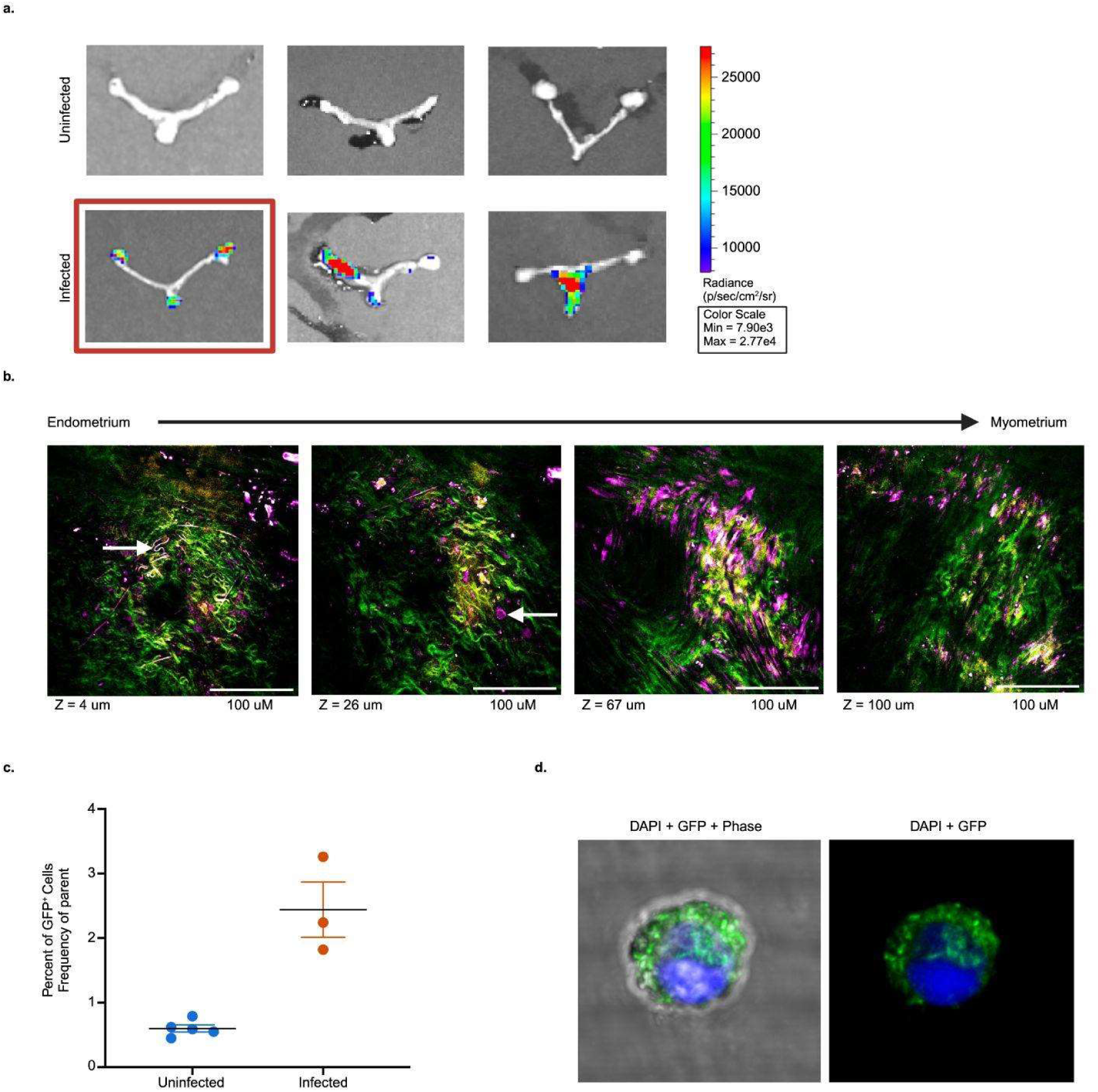
*B. burgdorferi* is present in the reproductive tract of infected mice. A. Representative *In Vivo* Imaging System (IVIS) images of female C57BL/6 (B6) murine reproductive tract tissue intradermally infected (10^5^ bacteria) with ML23 Luc-*B. Burgdorferi* for 9 weeks. Images were normalized across all cages for better visualization with a set radiance (7.90 × 10^3^ to 2.77 × 10^4^ p/sec/cm^2^/sr). B. Nonlinear imaging of Luc-*B. burgdorferi* infected uterine tissue from red-boxed Figure 1A spanning from endometrium to myometrium with Z=4-100 uM. Collagen is represented in green and lipid interface is represented in purple. C. Percent of GFP^+^ reproductive tract tissue digested cells of B6 mice that were uninfected (N=5) versus infected with GFP-*B. burgdorferi* (N=3), 12-months post-intraperitoneal injection with GFP-*B. burgdorferi*. Baseline was set such that GFP positivity in uninfected mice is under 1%. D. Microscopy of fluorescence activated cell sorted GFP^+^ reproductive tract tissue digestion cells from female B6 mice infected with GFP-*B. burgdorferi*, 15-months post-intraperitoneal injection with GFP-*B. burgdorferi*. DAPI is represented in blue and GFP is represented in green.

The gynecologic pathology from GFP-*B. burgdorferi* 14-month infected B6 female mice was examined macroscopically post-sacrifice but prior to tissue harvesting. After an incision to open the peritoneum of the infected mice is made to access the reproductive tract, elements of uterine pathology were often observed upon gross examination at necropsy. Images of infected murine reproductive tracts, compared to that of aged matched uninfected controls, show gross pathology that often includes enlargement of the uterus, ovarian cysts, yellowed tissues, as well as occasional torsion of uterine horns (Figure 2B).

**Figure 2.**
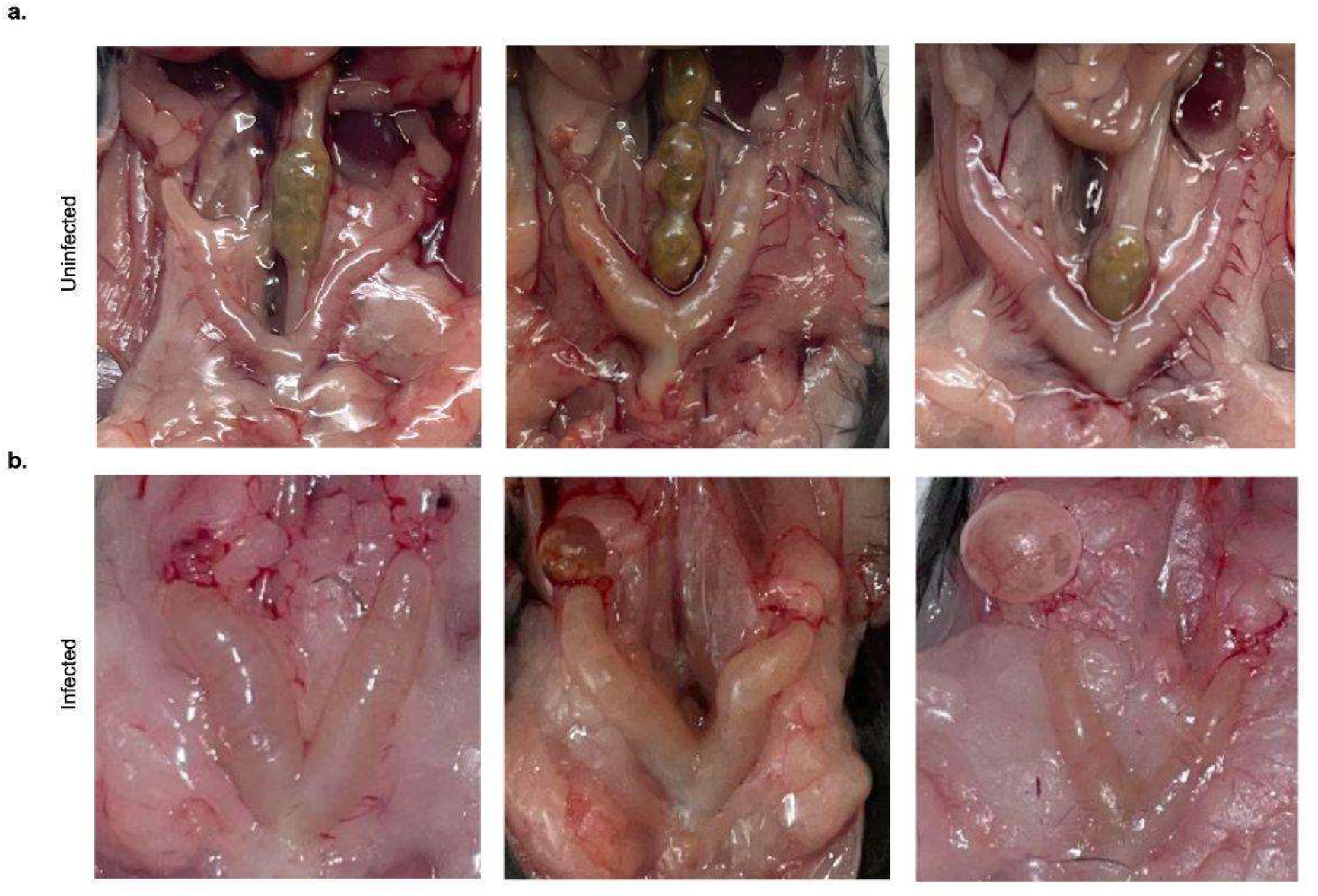
Gross pathology of GFP-*B. burgdorferi* infected mice. Gross pathology images of murine female reproductive tracts from A) uninfected 12-month old B6 female mice compared to that of B) infected B6 mice 14-months post-intraperitoneal injection of 10^5^ GFP-*B. burgdorferi*, infected at 3 months of age. N=3 of 4 uninfected and N=3 of 9 infected shown.

C3H/HeJ (C3H) mice, which develop multiple symptomatic similarities to humans with *B. burgdorferi* infection, were also studied (Extended Data Figure 2). Even though C3H female reproductive tract pathology also worsens with established 12-month GFP-*B. burgdorferi* infection, like B6 females in Figure 2B, uninfected C3H animals often develop ovarian cysts and uterine pathology with age, regardless of infection, making C3H female long-term infection and aged mice more difficult to study compared to that of B6 female mice (Extended Data Figure 2). Therefore, B6 mice were primarily studied in this LD gynecologic pathology infection model.

**Extended Data Figure 2.**
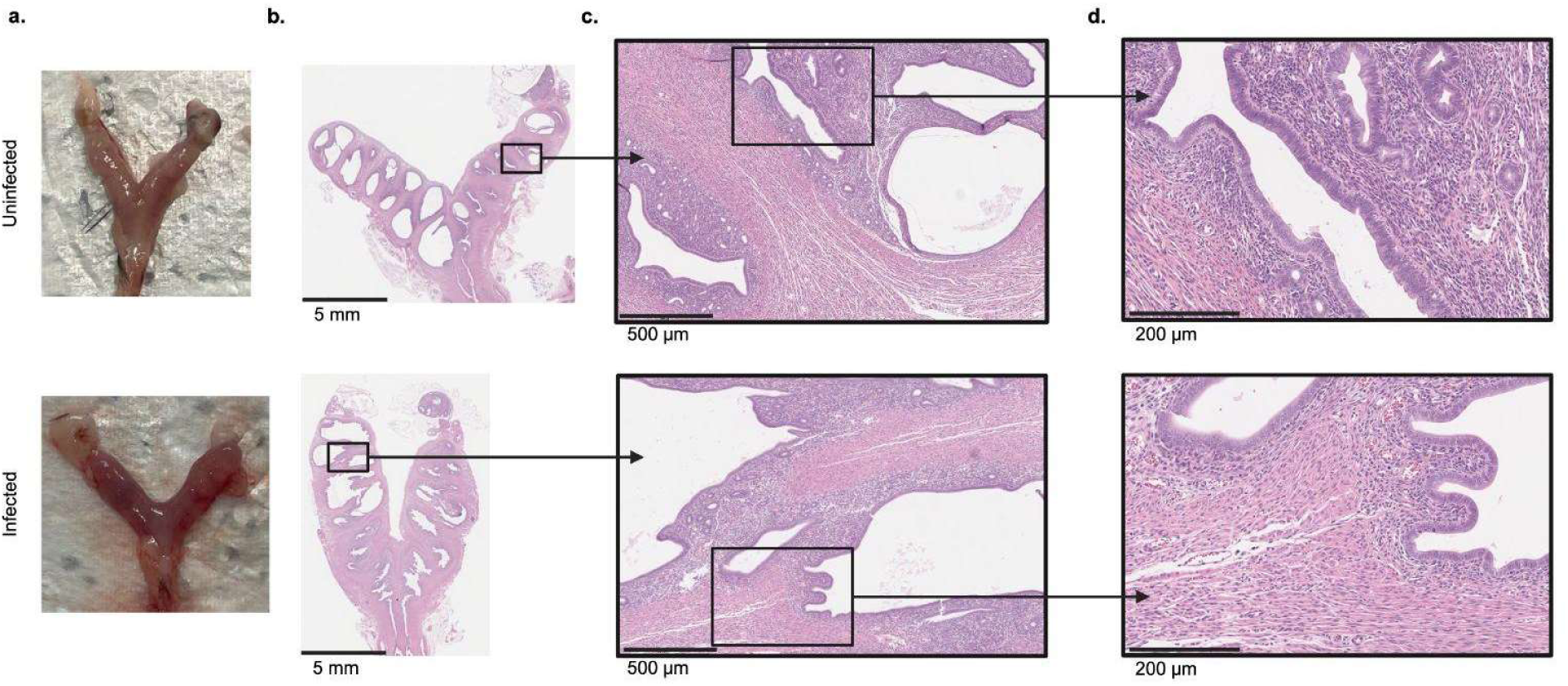
Gross pathology of GFP-*B. Burgdorferi* infected mice. A. Gross pathology images of murine female reproductive tracts from C3H/HeJ (C3H) mice 12-months post-injection that were either uninfected or infected intraperitoneally with 10^5^ GFP-*B. burgdorferi*. Histology of murine female reproductive tracts from C3H mice from panel A at different magnifications where scale bar = 5mm (panel A), 500 μm (panel C), and 200 μm (panel D). N=1 out of 5 representative reproductive tracts sectioned longitudinally.

### Gynecological pathologies of the infected murine uterus and vagina

The female reproductive tract (including ovaries, fallopian tubes, uterus, cervix and vaginal canal) of representative GFP-*B. burgdorferi* 12-month infected female B6 mice as well as controls’ gross pathology images are shown (Figure 3A). Organs sectioned longitudinally down the length of the fixed tissue (from ovaries down to cervix) were stained for hematoxylin and eosin (H&E) for histology. Histology images show that the infected organs have an increase in the diameter of the uterine horns at their largest width (2.79 mm vs 2.29 mm), which corroborate that of the gross pathology results shown in Figure 2 (Figure 3B). Glandular cysts and endometrial hyperplasia are seen in infected uterine tissue (Figure 3C & Figure 3D). The vaginal canal was sectioned longitudinally for one specimen and transversally for the remainder of the infected B6 murine cohort. For all of the infected B6 animals, histology images show signs of inflammation in the upper (closest to cervix) and middle thirds of the vaginal canal, including epithelial hyperplasia, polymorphonuclear and mononuclear cell infiltration of the epithelium, and extensive epithelial desquamation covering the canal’s lumen (Figures 3f-h). Additionally, researchers often observed an increase in discharge of infected mice (data not shown). This vaginal inflammation also correlates with external genitalia soft tissue swelling (Figure 3E).

**Figure 3.**
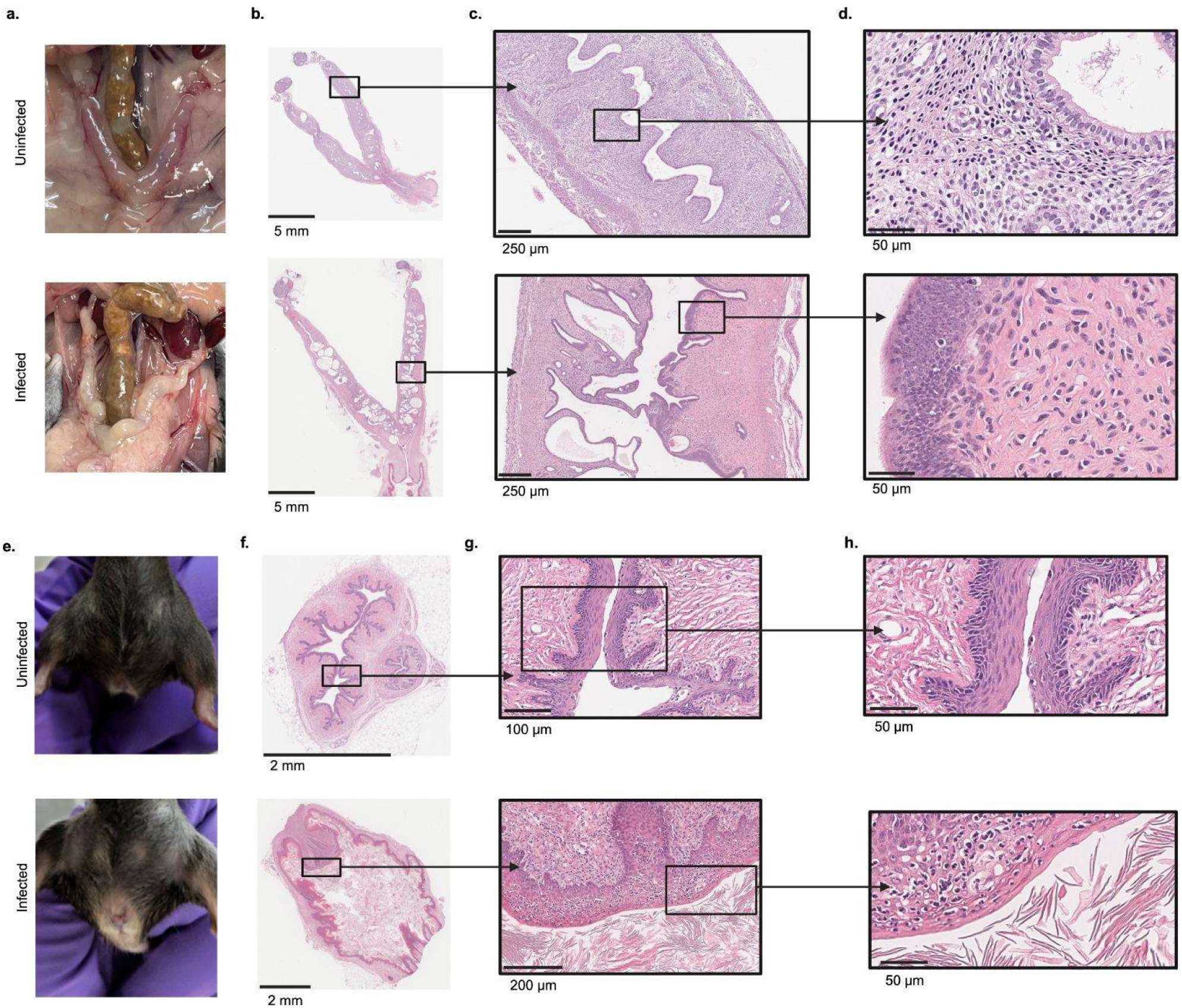
Gynecological pathologies of the infected murine uterus and vagina. A. Gross pathology images of representative murine female reproductive tracts from B6 mice 12-months post-injection that were either uninfected or infected intraperitoneally with 10^5^ GFP-*B. burgdorferi* at 8-weeks of age. Hematoxylin and eosin (H&E) histology of murine female reproductive tracts sectioned longitudinally from B6 mice shown in panel A, at different magnifications where scale bar = 5mm (panel B), 250 um (panel C), and 50 um (panel D). E. Representative images of the external genitalia of *B. burgdorferi*-infected and uninfected female aged, reproductively senescent and breeding-naïve B6 mice. Transversal sections of the vagina at different magnifications (F=2 mm, G=100 µm (top) and 200 µm (bottom), H=50 µm). N=1 out of 5 uninfected and N=1 out of 3 infected representative uteri and vaginas are shown.

### *B. burgdorferi*-induced gynecological pathologies worsen with increased age of female mice at time of infection

The immunological host response to infection has been known to vary with age and sex across a variety of pathogens^24–29^. Since mice from Figure 2 were left infected for 12 months and euthanized at 14 months of age and mice from Figure 3 were left infected for 14 months and euthanized at 17 months of age, we were interested in investigating if age of mice with *B. burgdorferi* infection or total length of infection time contributes to the infection model’s gynecologic pathology. B6 female mice were infected with Luc-*B. burgorferi* at either 15-weeks old (“young”, N=5) or 1-year old (“aged”, N=4) and euthanized after 9-weeks of infection. Female mice at 15-weeks old are typically sexually mature whereas at 1-year old they become reproductively senescent^30^. IVIS imaging (Figure 4A) & quantification compared to organ weight (Figure 4B) show that 3 of the N=4 infected aged murine organs had higher flux (photons/sec/cm^2^/steradian) than that of any of the infected young murine organs, which all had similar total flux to that of the uninfected animals’ organs. The gross pathology (Figure 4C) of the fixed, harvested reproductive tracts showed more detrimental impact of infection with increased age at time of infection, showing an increase in uterine horn size, an increase in cysts, and occasional torsion of the uterine horns. Histology of the two reproductive tracts with the largest bacterial burden measured by total flux values and circled in figure 4B show disfigured tissue with endometrial hyperplasia and glandular cysts (Figure 4D). These data suggest that age may impact the extent of uterine pathology with infection beyond the length of time infected. While 15-week old mice did not develop overt gynecological pathologies within 9-weeks of *B. burgdorferi* infection, their 1-year old counterparts did. One-year old females display *B. burgdorferi* infection-associated morbid gynecologic pathology either when infected young and left infected for 12-months, or when infected at 12-months old with 9-weeks of infection.

**Figure 4.**
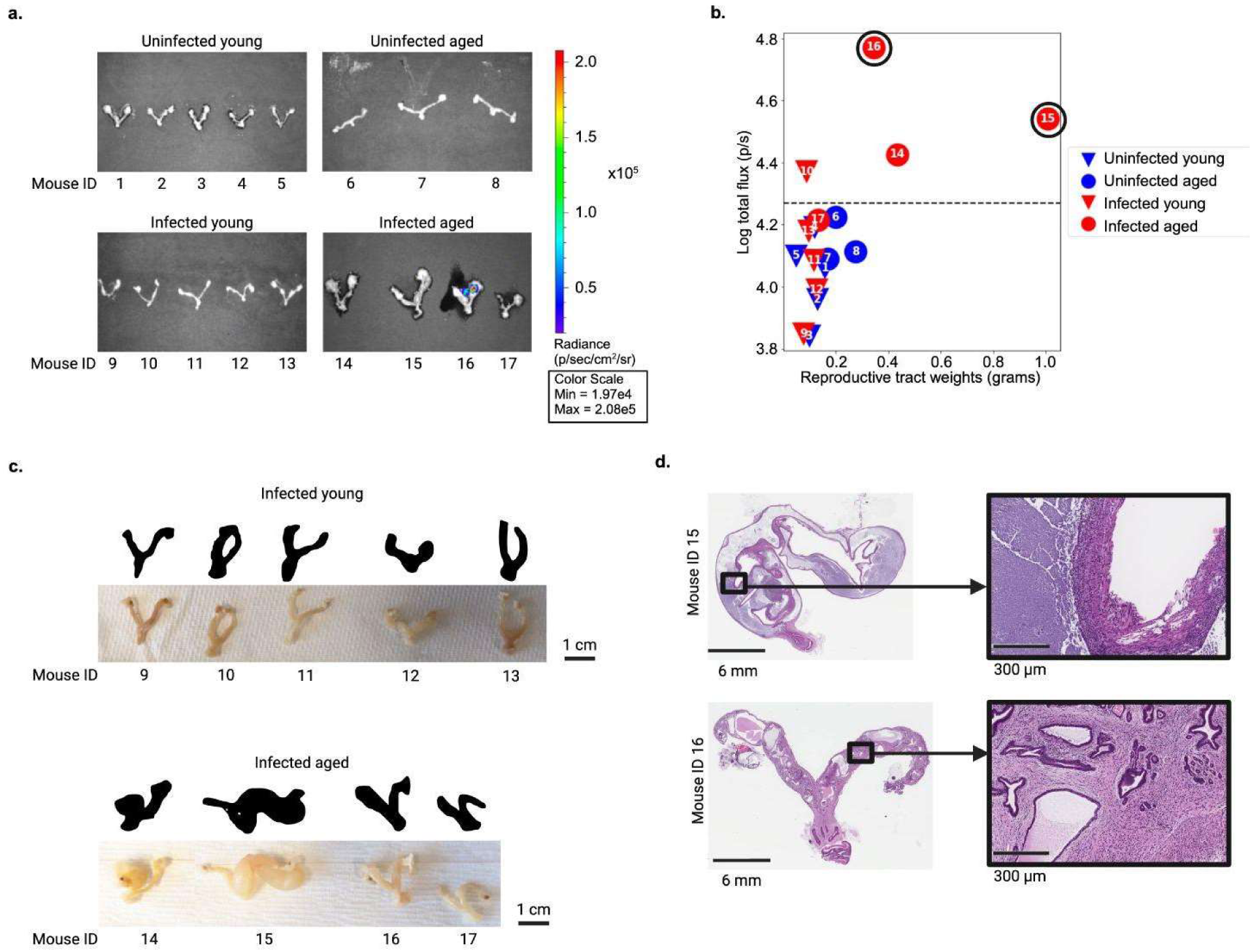
*B. burgdorferi*-induced gynecological pathologies worsen with increased age of female mice at time of infection. A. *In Vivo* Imaging System (IVIS) images of reproductive tract (ovaries, uterus, cervix, etc.) tissue from B6 mice that were uninfected or infected for 9-weeks with 10^5^ N40 Luc-*B. Burgdorferi* intradermally infected at either 15-weeks old (“young”) or 1-year old (“aged”). B. Reproductive tract weights were compared to IVIS total flux quantification for each mouse’s reproductive tract as shown in panel A. The total flux was plotted on a logarithmic scale. The two mice with the highest reproductive tract flux are circled. C. Gross pathology images and silhouettes of female reproductive tracts harvested from infected young and infected aged mice. D. Histology of murine female reproductive tracts with the two highest luc-*B. burgdorferi* flux as measured in panel B (mice circled in 4B) at different magnifications where scale bar = 6 mm & 300 um. Reproductive tracts were sectioned longitudinally down the length of the tissue.

### Human epidemiological evidence of gynecological pathologies

We sought to further analyze whether LD history increased risk of adverse gynecological outcomes in humans. Using the All of Us Research Program, we found that female individuals diagnosed with LD had increased rates of gynecological complications when adjusted for demographics, location, and relevant risk factors using logistic regression (Figure 5B), including dysmenorrhea (1.56-fold odds ratio, p = 0.002), endometriosis (1.93-fold odds ratio, p = 3.4e-5), uterine fibroids (1.42-fold odds ratio, p = 8.8e-4), menorrhagia (1.54-fold odds ratio, p = 0.016), and miscarriage (1.61-fold odds ratio, p = 0.039). The association remained, albeit diminished in effect, when adjusting for doxycycline administration following Lyme diagnosis for dysmenorrhea (1.62-fold odds ratio, p = 0.02), endometriosis (1.94-odds ratio, p = 0.0047), uterine fibroids (1.71-odds ratio, p = 1.28e-4), and menorrhagia (2.03-odds ratio, p = 0.0089, details in Supplementary Material). This striking effect was replicated in Truveta, a large electronic healthcare record aggregation database derived from ∼110M individuals, where odds of dysmenorrhea (1.69-fold hazard ratio, p = 1.74e-17), endometriosis (1.195-fold hazard ratio, p = 0.0171), uterine fibroids (1.242-fold hazard ratio, p = 0.0019), and menorrhagia (1.355-fold hazard ratio, p = 1.14e-12) were all increased in incidence in females with LD compared to females without this disease (see Supplementary Material). In addition, diagnosis with Lyme resulted in incident cases, supporting a causal relationship (Figure 5C). Finally, we corroborated these findings in Finngen (Figure 5F), where we also observed higher rates of Transvaginal ultrasound examination of uterus, parametria and lower abdomen (LC2BE, OR 1.4, p = 1.9e-9), Ultrasound examination of uterus and parametria (LC2AE, OR 1.6, p = 3.2e-9), and Vaginal administration of clindamycin (G01AA10, OR 1.4, -log10p = 4.0e-7) among LD cases.

**Figure 5.**
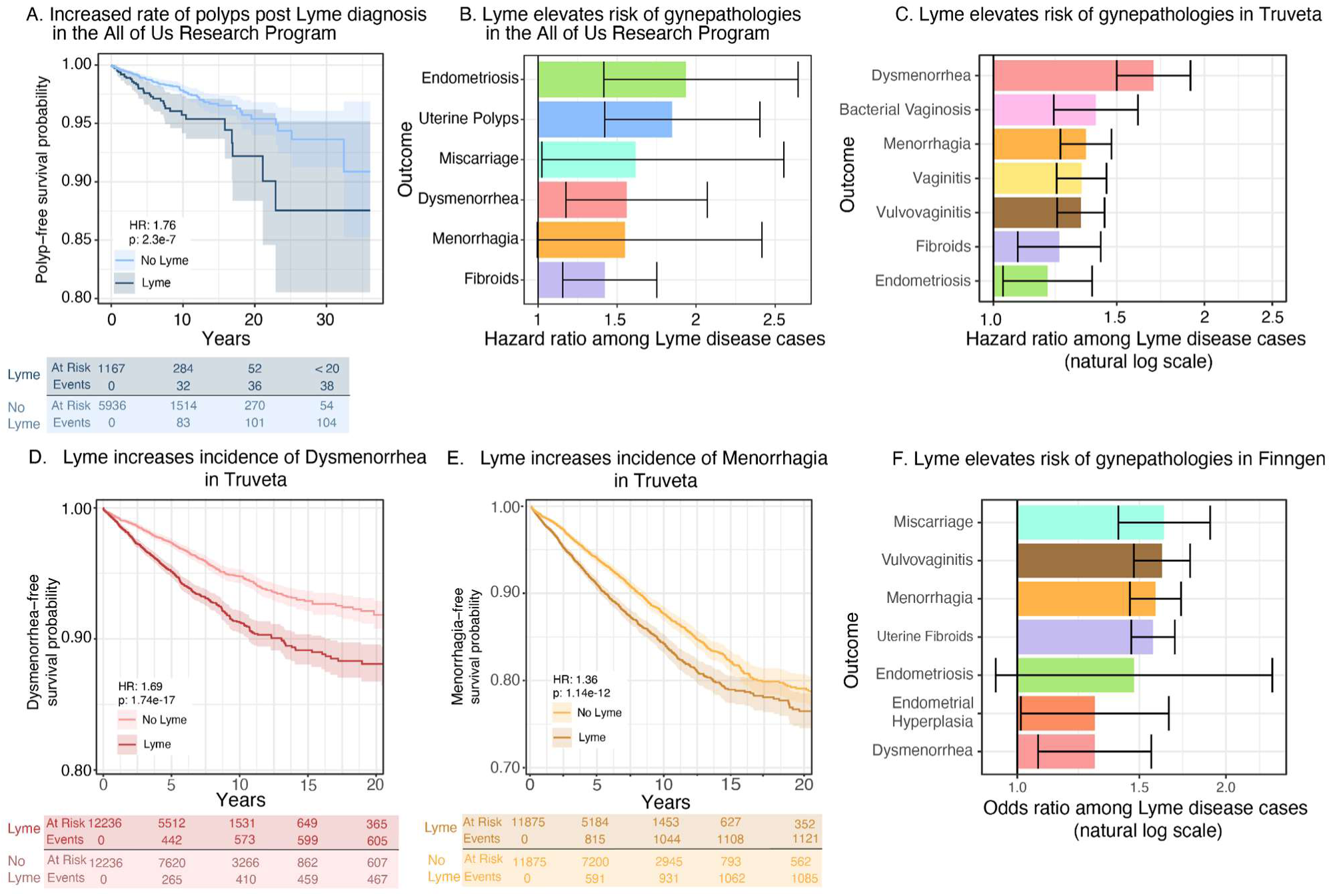
Human epidemiological evidence of gynecological pathologies. A. Uterine polyps are increased in incidence following Lyme diagnosis in the All of Us Research Program. Additional disease outcomes are presented in the Supplementary Material. B. Incidence of numerous gynecological pathologies, including endometriosis, uterine polyps, miscarriage, and dysmenorrhea, are significantly increased following diagnosis with LD. C. The association of gynecological pathology outcomes with LD is elevated in an independent replication cohort, Truveta. D. Kaplan-Meier curve showing survival time varies in an LD-dependent fashion and that LD elevates risk of Dysmenorrhea. E. Kaplan-Meier curve showing survival time varies in an LD-dependent fashion and that LD elevates risk of menorrhagia. F. The odds ratio of gynecological pathology outcomes is elevated among individuals with LD in an independent replication cohort, Finngen.

As a result of observing an age-dependent effect in mice, we then examined whether individuals recruited before age 45 or after age 60 in the All of Us Research Program^31^ had different effect sizes. We observed moderately elevated odds ratios for multiple gynecological pathologies among individuals recruited at an older age in All of Us (Extended Data Figure 3A). Truveta analysis also indicated a trend towards elevated risk (Extended Data Figure 3B), including associated with developing dysmenorrhea in 45 year old individuals with history of Lyme Disease (HR: 1.80) compared to 15 year old individuals (HR: 1.44, difference z-test p = 0.195, Extended Data Figure 3C-D). The effect of LD on other outcomes were relatively constant with age, including for vulvovaginitis (Extended Data Figure 3E-G).

**Extended Data Figure 3.**
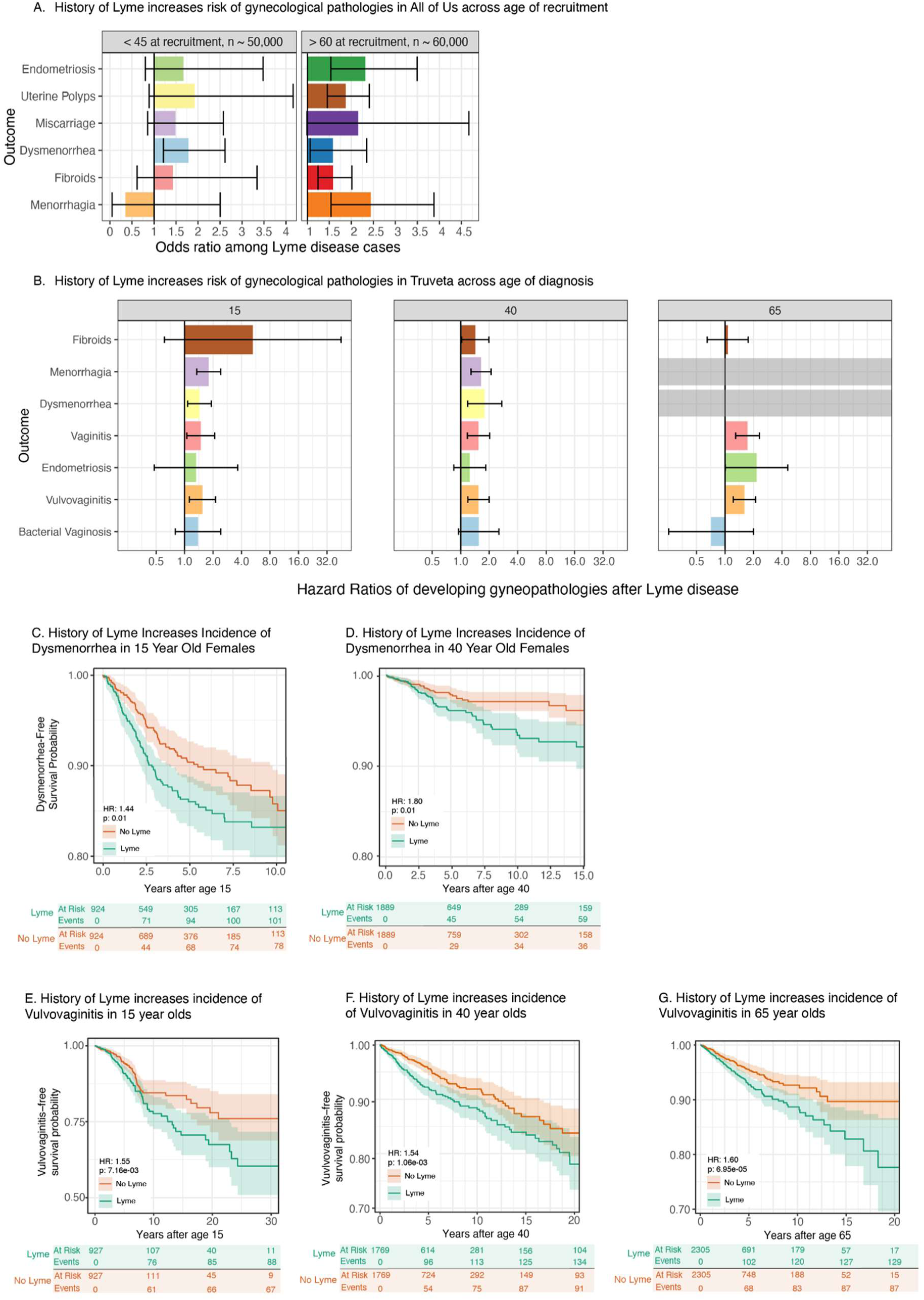
Age dependence in humans. A. Comparing individuals in All of Us recruited below age 45 versus those recruited above age 60, there is a trend toward elevated EHR-recorded gynecological pathologies among participants recruited at an older age. B. This effect was consistent with our tests of age dependence in Truveta, where baseline ages of 15, 40, and 65 showed similar trends for a broad range of gynecological pathologies. Note that incidence of menorrhagia and dysmenorrhea was not tested in individuals over 65 years of age due to diagnostic criteria. C. Kaplan-Meier curve showing survival time varies in an LD-dependent fashion and that LD elevates risk of dysmenorrhea in individuals with a baseline time starting at age 15. D. Kaplan-Meier curve showing survival time varies in an LD-dependent fashion and that LD elevates risk of dysmenorrhea in individuals with a baseline time starting at age 40. E. Kaplan-Meier curve showing survival time varies in an LD-dependent fashion and that LD elevates risk of vulvovaginitis in individuals with a baseline time starting at age 15. F. Kaplan-Meier curve showing survival time varies in an LD-dependent fashion and that LD elevates risk of vulvovaginitis in individuals with a baseline time starting at age 40. G. Kaplan-Meier curve showing survival time varies in an LD-dependent fashion and that LD elevates risk of vulvovaginitis in individuals with a baseline time starting at age 65.

Overall, these findings suggest a strong and repeated association between LD and incidence of gynecological outcomes across ages, supporting the notion that uterine health is partially compromised by *B. burgdorferi* infection.

## Discussion

It is currently unappreciated that LD can lead to gynecological pathologies. This is the first murine study to demonstrate that *B. burgdorferi* infection can directly cause extensive gynecologic pathology. It is also the first population-based study to demonstrate a strong association between LD and subsequent gynecologic conditions. Our data suggest that LD may have an additional manifestation as a gynecological disease. Previous rodent studies have shown that *B. burgdorferi* can be regularly isolated from the bladder of infected white-footed mice^32^ as well as the uterus of female C3H/HeN mice upon infection-pregnancy studies^20^. Previous human studies have shown that *B. burgdorferi* can be cultured from LD patients’ genital secretions^33^ and was associated with erosive vulvovaginitis in a case study^34^. In addition to finding metabolically active *B. burgdorferi* in the reproductive tract of long-term infected breeding-naïve female mice, we also identify an association with increased gynecological pathologies. These are observable via gross pathology, including disfiguring enlargement of the uterus, ovarian cysts, yellowed tissues, external genitalia swelling, as well as occasional torsion of uterine horns. Additionally, histology of the uterus displays glandular cysts and endometrial hyperplasia, and the pathological findings are not limited to the uterus and cervix; in all infected specimens, the vagina exhibited histological evidence of inflammation associated with infection as well. The presence of mononuclear and polymorphonuclear infiltration suggests that this inflammatory process might not be an isolated event, but a recurring pattern.

Previous human and murine research has established associations between acute infection or infection-associated chronic illnesses and gynecological conditions. A population-based cohort study of 79,512 patients with inflammatory diseases of the cervix, vagina, and vulva found that compared to age-and-sex matched controls, lower genital tract infection was associated with a substantially elevated risk for developing endometriosis regardless of comorbidities^35^. Women with Myalgic Encephalomyelitis/Chronic Fatigue Syndrome (ME/CFS), an infection-associated chronic illness, have elevated rates of endometriosis, Polycystic Ovary Syndrome (PCOS), and uterine fibroids^36,37^. Heavy menstrual bleeding is commonly experienced by female patients with Long COVID or ME/CFS^36^. Muraoka et al. found fusobacterium in 64% of endometrial tissue samples from women with endometriosis compared to 7% of controls, with higher frequencies of fusobacterium in those with endometriosis. Fusobacterium infected female mice developed endometriotic lesions, which were reduced with antibiotic treatment^38^.

Biological mechanisms of endometriosis are not well understood^39^. More research is critically needed on the relationship between infection, inflammatory responses to infection, and the development of gynecological pathologies, including endometriosis. Khan’s “bacterial contamination hypothesis” proposes that microbes may contribute to endometriosis growth and progression via bacterial recognition pathways such as LPS/TLR4 and peptidoglycan/TLR2 signaling cascades^38,40–42^. TLR2 and TLR1/2 heterodimers have been shown to be the main recognition receptors of *B. burgdorferi*^43–46^. In the context of the bacterial contamination hypothesis of endometriosis, these inflammatory cascades may offer a potential mechanism to explain the increased risk of endometriosis following LD. Proinflammatory stimuli are known to decrease expression of progesterone receptors, which are crucial for pregnancy, and may be involved in endometriosis-associated infertility^47,48^. This increased inflammatory state of the female reproductive tract may also offer a potential mechanism for increased risk of dysmenorrhea and miscarriage.

Our findings of inflammation in the vagina and vulva of infected specimens suggest a link between *B. burgdorferi* infection and direct or indirect damage to the genital organs. While the route of transmission for genital infection remains a matter of speculation, the evidence of vulvar and vaginal inflammation in our model, combined with previous studies, suggests that *B. burgdorferi* infection alters the integrity of the female genital tract.

Importantly, we find *B. burgdorferi*-induced gynecological pathologies worsen with age, particularly in reproductively senescent female mice. C57BL/6 mice infected with *B. burgdorferi* have generally been shown to demonstrate only mild disease manifestations, which makes the extent of gynecological disease observed in older female animals with *B. burgdorferi* infection even more surprising. We find that either long-term (12+ months) untreated *B. burgdorferi* infection and short-term (9-weeks) infection of 1-year aged mice both lead to worse pathologies than that of 8-15-week old mice infected short-term (9-weeks). These data suggest the importance of understanding the full extent of LD’s impact on the reproductive tract throughout life and hormonal phase which could be contributing to the age dependency of increased gynecologic pathology. Sex differences in the immune response to infection, especially in regard to reproductive health, are understudied^49,50^. Therefore, we suggest that future studies investigate whether *B. burgdorferi* infection may colonize the male reproductive organs or if infection is associated with reproductive pathologies in males. Since age potentially has an impact on LD gynecologic pathology and “hormones act as modulators of the host reaction against trauma and infection”^51^, we suggest that future studies investigate the impact of hormones and hormonal dysregulation on this infection model, as well as any potential therapeutic impact of hormonal interventions. Furthermore, future studies should also investigate antibiotic treatment of *B. burgdorferi* infection in the context of gynecological diseases, as well as the importance of hormone levels, age, and cycling on gynecological pathological sequelae. We demonstrate direct evidence of *B. burgdorferi* infection in the female murine reproductive tract and association with increased downstream risk of gynecological pathologies in both mice and humans (Figure 6), highlighting the importance of future investigation in how *B. burgdorferi* infection drives gynecological disease.

**Figure 6.**
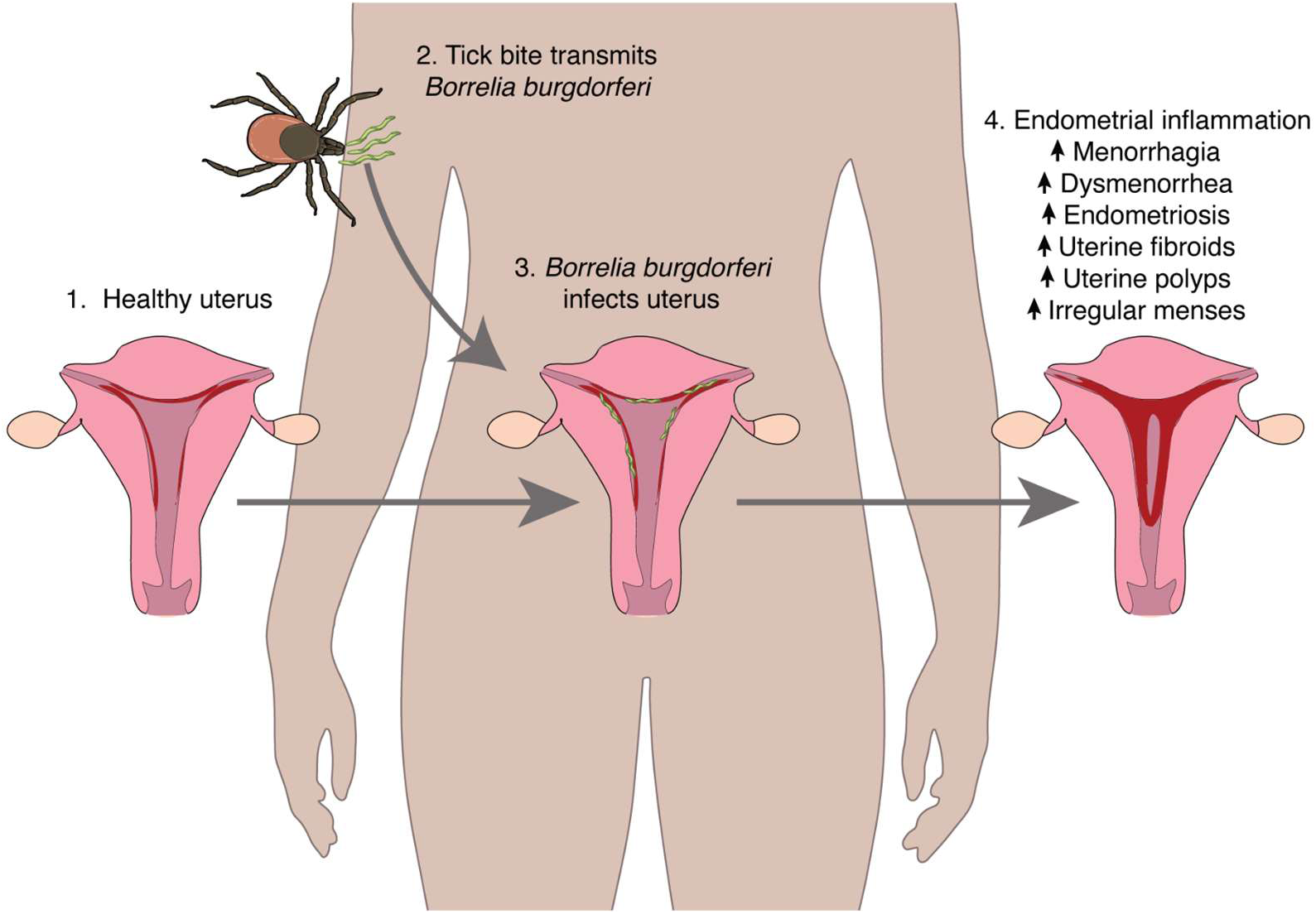
Overall model of gynecological pathology development following LD exposure.

## Methods

### Truveta

Clinical data was compiled from the Truveta database and consisted of deidentified medical records from 30 health care organizations representing ambulatory centers, hospitals, imaging centers, and clinics and medical offices. Truveta provides access to continuously updated and linked EHR and claims data including demographics, diagnoses, encounters, immunizations, medications, laboratory results, procedures, clinical notes, and images. Truveta Data used in this study was accessed in June and October 2024. Through syntactic normalization, similar data fields from different health care organizations are mapped to a common schema referred to as the Truveta Data Model (TDM). Once organized into common fields, the values are normalized to common ontologies such as ICD-10, SNOMED-CT, LOINC, RxNorm, and CVX, through semantic normalization. The normalization process employs an expert-led, artificial intelligence driven process to accomplish high-confidence modeling at scale. De-identification is attested to through expert determination in accordance with the HIPAA Privacy Rule. This study used only de-identified patient records and was approved under IRB FHIRB0011190 at Fred Hutchinson Cancer Center.

### All of Us analysis

Clinical and survey data from All of Us^31^ was aggregated for all female participants with EHR records available. Concept codes were used for diagnoses as follows:

~~~
Uterine fibroids: standard_concept_name **==** “Uterine leiomyoma“
Miscarriage: standard_concept_name **==** “Miscarriage without complication”
Endometriosis: standard_concept_name **==** “Endometriosis (clinical)“
Dysmenorrhea: standard_concept_name **==** “Dysmenorrhea“
Cervicitis: standard_concept_name **==** “Cervicitis and endocervicitis“
Inflammatory disease of the uterus: standard_concept_name **==** “Inflammatory disease of the uterus“
Pelvic inflammatory disease: standard_concept_name **==** “Female pelvic inflammatory disease“
Irregular menstruation: standard_concept_name **==** “Irregular periods” Excessive and frequent menstruation: standard_concept_name **==** “Excessive and frequent menstruation“
Uterine polyps: standard_concept_name == “Polyp of corpus uteri” Menorrhagia: standard_concept_name **==** “Menorrhagia“
Bacterial vaginosis: standard_concept_name **==** “Bacterial vaginosis“
~~~

In addition, we defined simplified gender identities (Man, Woman, Other), ethnicities (Hispanic or Latino, Not Hispanic or Latino, Other), and races (Black or African American, White, Asian, Middle Eastern or North African, Native Hawaiian or Pacific Islander, Multiple, Other). Individuals’ input time since birth was defined as the difference in years between January 1, 2023 and their birthday (as the 6-month mark after end of censorship).

Individuals were matched using MatchIt for propensity score matching, with 20 controls chosen for every case individual. The exact matching command is available in the supplemental materials, along with plots of the balance pre-versus post-matching. For all controls, the date of Lyme diagnosis for the corresponding case was set as the start of follow-up. Individuals who died before this date (n < 20 controls out of ∼25,000) were censored at time 0. For all other individuals, right censor time was defined as midnight on July 1, 2022, as this was empirically determined to be the right censor time for all EHR data in Release 7 used here.

MatchIt was conducted as follows:

~~~
match **<-** matchit(lyme **∼** age **+** state **+** gender.simple **+** race.simple **+** ethnicity.simple **+** high_school_education **+** median_income **+** poverty **+** no_health_insurance, data **=** person.match, method **=** “nearest”, distance **=** “glm”, ratio **=** 20)
~~~

The following cox proportional hazard model was used for the “complex” main analysis:

~~~
cox **<-** coxph(Surv(survTime, endpoint) **∼** lyme **+** surveymenopause **+** surveyht **+** surveyob **+** state **+** high_school_education **+** gender.simple **+** rural **+** race.simple **+** ethnicity.simple **+** age **+** median_income **+** poverty **+** no_health_insurance
~~~

Where high_school_education, median_income, poverty, and no_health_insurance are the fraction of individuals in the zip code who completed high school education, their median income, are below the poverty line, and do not have health insurance, respectively. Rural is defined based on the “Delayed Medical Care: Rural Area” survey question. surveymenopause is defined as the answer to the “Menstrual Stopped: Yes None” from the overall health survey. surveyht and surveyob are self-reported hypertension and obesity from the personal and family health survey, respectively. State is defined as the first digit (region) of the individuals’ reported zip code.

The following cox proportional hazard model was used for the “simple” sensitivity analysis:

~~~
cox **<-** coxph(Surv(survTime, endpoint) **∼** lyme **+** age **+** race.simple **+** rural **+** ethnicity.simple **+** state, data**=**data)
~~~

The following cox proportional hazard model was used for the doxycycline interaction sensitivity analysis:

~~~
cox **<-** coxph(Surv(survTime, endpoint) **∼** lyme **\*** doxy **+** age **+** race.simple **+** rural **+** ethnicity.simple **+** state, data**=**data)
~~~

Additional details and plots are available in the Supplementary Materials.

### FinnGen analysis

FinnGen is a public-private partnership registry-based study of Finnish residents combining genetic and electronic health record data from different registers, for example, primary care and hospital in- and out-patient visits. We used data from FinnGen release 12 (R12) that contains data on up to 500,348 participants of Finnish ancestry from newborns to the age of 104 at baseline recruitment^52^. The diagnosis of Lyme disease was based on ICD-codes (ICD−10: A69.2, ICD-9: 1048 A), which were obtained from the Finnish national hospital and primary care registries.

We utilized logistic regression analysis between diagnosis of Lyme disease and Menorrhagia, Miscarriage, Endometrial hyperplasia, Endometriosis, Uterine fibroids, Vulvovaginitis, Dysmenorrhea and adjusted the analysis for age and population structure. The analyses were run with R studio version 4.4.2 using the *glm* package in R.

### B. burgdorferi culture

In a sterile biosafety cabinet, stock vials of *B. burgdorferi* (1×10^7^ *B. burgdorferi*/tube), stored at - 80°C, were thawed into BSK-H complete medium with 6% rabbit serum (50 mL, Sigma-Aldrich) or BSK-II complete medium with 6% rabbit serum (50 mL, generously made by the Chou lab or made in-house following protocol from Jenny Hyde’s Lab). Strain *B. burgdorferi* B31 A3 GFP was generously provided by George Chaconas. ML23-pBBE22luc was generously provided by Jenny Hyde^53,54^. N40D10/E9 (N40) was generously provided by Nikhat Parveen^55^. Each culture was in a sealed tube incubated at 37°C for 7 days, without a water bath, at a pH of 7.6, unless otherwise stated.

### Fluorescence-Activated Cell Sorting (FACS) of *B. burgdorferi* cultures

The concentration, viability of cells and percentage of GFP expression was calculated with Fluorescence-Activated Cell Sorting (FACS). At Stanford University School of Medicine FACS Core, FACS was conducted on a BD LSRFortessa with BD FACS Diva software. For FACS analysis of *B. burgdorferi* at Stanford, LSRFortessa cytometer threshold levels were modified to parameter SSC 400 and voltages were set to FSC 300 and SSC 230, collected in log mode. At Massachusetts Institute of Technology Koch Institute Flow Cytometry Core, FACS was conducted on either a BD Symphony A3 HTS I or a BD LSRFortessa HTS II. For FACS analysis of *B. burgdorferi* at MIT, Symphony cytometer threshold levels were modified to parameter SSC 400 and voltages were set to FSC 250 and SSC 240, collected in log mode. For FACS analysis of *B. burgdorferi* at MIT, LSRFortessa cytometer threshold levels were modified to SSC 800 and voltages were set to FSC 300 and SSC 240. *B. burgdorferi* were counted and assessed for integrity via FACS but not assessed for motility.

### Murine B31 A3 *B. burgdorferi*-GFP infection

Bacterial concentration was determined by flow cytometry (BD LSRFortessa, Stanford University Flow Core). Mice were anesthetized by 3% isoflurane inhalation and injected intraperitoneally with either 10^5^ GFP-*B. burgdorferi* from a 7-day culture resuspended in PBS or PBS alone (if “uninfected”).

Female B6 (N=5 uninfected aged 8-weeks, N=3 infected aged 8-weeks) and C3H/HeJ (C3H) (N=5 uninfected aged 8-weeks, N=5 infected aged 8-weeks) mice were left infected for 1-year and used for flow cytometry of single-cell female reproductive tract tissue digestions, reproductive tract gross pathology imaging, and reproductive tract histology.

Female C57BL/6 (B6) (N=6 infected aged 8-weeks) mice were left infected for 15-months and used for reproductive tract digestion, Fluorescence Activated Cell Sorting (FACS), and fluorescence microscopy.

Female B6 mice (N=9 infected aged 3-months) were left infected for 14-months and used for downstream gross pathology imaging. Female B6 mice (N=4 uninfected aged 12-months) were compared to infected and used for downstream gross pathology imaging.

All mice were acquired from Jackson Laboratories (Bar Harbor, ME).

### Murine *B. burgdorferi*-luciferase infection

N40D10/E9 (N40) and ML23 Luciferase *B. burgdorferi* were cultured in BSKII media and 6% rabbit serum (Millipore Sigma) at a pH of 7.6 in 37°C for 5 days. ML23 cultures also contained a selection antibiotic, Kanamycin. Bacterial concentration was determined by flow cytometry (Becton Dickinson LSR-II, Koch Flow Core). The cultured *B. burgdorferi* were resuspended in 500 μL of 0.2% uninfected B6 murine serum (diluted in PBS). Female C57BL/6 mice (n= 5 per condition) aged 6-weeks or 15-weeks and 1-year (ages at time of infection stated in figure legends) were anesthetized by 3% isoflurane inhalation and injected intradermally with 10^5^ *B. burgdorferi* (50 μL volume) per mouse.

All mice were acquired from Jackson Laboratories (Bar Harbor, ME).

### Confirmation of murine *B. burgdorferi*-luciferase infection using In Vivo Imaging System (IVIS)

To confirm infection with a luciferase strain of bacteria, an injection of 100 μL sterile filtered D-luciferin (GoldBio) reconstituted in sterile PBS at 277 mg/kg was performed intraperitoneally. The hair on the backs of the mice was removed using an electric shaver with then the application of Nair™ for 30 seconds to 1 minute. Gauze wipes were used to clean the backs of the mice post-Nair™ in the following order: soaked in 70% ethanol, soaked distilled water, and a dry gauze. 15 minutes after the D-luciferin injection, mice were arranged in the In Vivo Imaging System (Perkin Elmer) then imaged for an exposure time of 1 minute.

### Murine *ex vivo* uterine *B. burgdorferi*-luciferase load imaging using In Vivo Imaging System (IVIS)

Reproductive tracts were cut through the vaginal canal and the upper portion of the reproductive tract (ovaries, uterus, cervix, etc.) were harvested from euthanized mice and immediately placed into PBS with luciferin (300 μg/mL) & 1 mM ATP solution for 15 minutes before imaging. Organs were then arranged in the In Vivo Imaging System (Perkin Elmer) and imaged for an exposure time of 1 minute. The reproductive organs were placed back into the PBS with luciferin (300 μg/mL) & 1 mM ATP solution before the 3D nonlinear imaging.

### Analysis of In Vivo Imaging System (IVIS) Data

All downstream IVIS analyses were performed, blinded with Living Image (Perkin Elmer). The average radiance values (photons/sec/cm²/sr) were achieved as the sum of radiance from each pixel within the rectangular gating of individual mouses’ organs. All images were normalized across all cages and time points with a set radiance range listed in the figure legend for visualization purposes.

### Nonlinear imaging of reproductive organs infected with *B. burgdorferi*-luciferase (ML23)

The female reproductive tracts were placed in PBS with luciferin (300 μg/mL) & 1 mM ATP solution for IVIS imaging prior to nonlinear imaging. The female reproductive tract with the highest level of flux via IVIS was selected for nonlinear imaging. 3D nonlinear imaging of living reproductive organs was achieved using a custom-built inverted nonlinear microscope^56,57^. A pulsed laser (Light Conversion, Cronus-3P) operating at 1300 nm was employed for simultaneous imaging of third harmonic generation (THG), three-photon fluorescence (3PF), and second harmonic generation (SHG). Signals were processed into images using custom-written LABVIEW acquisition software and visualized with Fiji software^58^. To perform 3D imaging of the reproductive organs, the uterine tissue was positioned with the endometrium side facing downward on the inverted microscope. The microscope stage (ASI, MS2000) was then controlled to shift the focal plane progressively deeper, from the endometrium (*Z* = 0 μm) to the myometrium.

### Murine ovarian and reproductive tract weight, gross pathology imaging, histology, and tissue digestion

To access the reproductive tract of euthanized mice, an incision was made to open the peritoneum. Photos of the reproductive tract organs in the peritoneum were captured. Gross pathology was recorded and photographed. Reproductive tracts were excised and immediately placed into either sterile PBS or 4% PFA (diluted to 4% from 16% PFA with PBS) (Beantown Chemical or Pierce, Thermo Scientific). Organs in 4% PFA for fixation were transferred to 70% ethanol after 24-hours and protected from light for long term storage. Prior to being weighed and imaged, fixed organs were blotted on paper towels to remove excess moisture as well as gently separated from any fat and excess non-uterine tissue. Organs were imaged with a ruler and cell phone camera and then weighed on a scale (Mettler AJ100). Fixed organs were sent to either HistoWiz (Brooklyn, NY) or the MIT Hope Babette Tang (1983) Histology Core Facility for histology. Fixed organs were sectioned longitudinally and transversally (from ovaries down to vagina) and stained with hematoxylin and eosin (H&E). Organs placed into sterile PBS were digested and dissociated in collagenase dissociation buffer at 30°C for 30 minutes, followed by trituration. Digested tissue was passed through a 70-μm cell strainer and washed in PBS. Single cell digestions of reproductive tract tissue were analyzed via flow cytometry within a few hours of digestion (please see Flow cytometry methods section below for more information).

### Flow cytometry, Fluorescence-Activated Cell Sorting (FACS), and imaging of reproductive tract cells with GFP-*B. burgdorferi* infection

Digested tissue was analyzed on a 350 Symphony flow cytometer or sorted on a FACS Aria (BD Biosciences) with BD FACS Diva software at Stanford University School of Medicine FACS Core. For fluorescent microscopy, fixed tissue and sorted cells were plated on glass slides, stained with DAPI and imaged by fluorescent confocal microscopy on Leica SP8 with whitelight laser inverted confocal (Leica) for the presence of GFP^+^ cells. Fluorescent microscopy was performed at the Beckman Imaging Core at Stanford (NIH S10 Shared Instrumentation Grant Award 1S10OD010580).

### Statistical analysis

All statistical methods and number of replicates are reported in figure legends. Error bars in all figures were calculated as standard error to the mean (SEM) in GraphPad Prism.

## Data Availability

All data presented in the current study are available upon reasonable request to the authors

## Acknowledgements

We would like to thank the ongoing support of the MIT Department of Comparative Medicine and the Koch Institute’s Robert A. Swanson (1969) Biotechnology Center for technical support, specifically Preclinical Imaging & Testing Core, for consultations and support for *in vivo* mouse experiments. We would also like to thank the Beckman Imaging Core at Stanford University, Stanford University School of Medicine FACS Core, as well as the Massachusetts Institute of Technology Koch Institute Flow Cytometry Core. This work has been supported by the transformational philanthropic support by the Emily and Malcolm Fairbairn donor advised fund. Imaging instruments used at the Beckman Imaging Core were supported by an S10 Shared Instrumentation Grant Award. N S-A, PP, and MWP are funded through a Microbiome Research Institute Pilot Award and the Klorfine Pilot Award. We would like to thank Anirudh Pai and Rachel Salomon for helping with large murine endpoint experiments. We would like to thank Thomas Donaghey for his support in lab management. We would also like to thank the members of the Tal Research Group, Dr. Nasa Sinnott-Armstrong’s laboratory, Dr. Linda Griffith’s laboratory, Dr. Doug Lauffenburger’s laboratory, Tal Raveh, Erin Sanders, and Sue Faber for helpful discussion. This work has been supported by the Instrumentarium Science Foundation (HMO).

## Abbreviations

LD: Lyme Disease
*B. burgdorferi*: *Borrelia burgdorferi*
EM: erythema migrans
IVIS: *In Vivo* Imaging System
luc-*B. burgdorferi*: Luciferase strain of *B. burgdorferi*
GFP-*B. burgdorferi*: Green Fluorescence Protein strain of *B. burgdorferi*
B6: C57BL/6J
C3H: C3H/HeJ
photons/sec/cm2/steradian: flux
H&E: hematoxylin and eosin
ME/CFS: Myalgic Encephalomyelitis/Chronic Fatigue Syndrome
PCOS: Polycystic Ovary Syndrome

## Figure creation

Text figure 1 (Created in BioRender. Tal, M. (2025) https://BioRender.com/i66j505), figure 2 (Created in BioRender. Tal, M. (2025) https://BioRender.com/l01b236), figure 3 (Created in BioRender. Tal, M. (2025) https://BioRender.com/b27v297), figure 4 (Created in BioRender. Tal, M. (2025) https://BioRender.com/h31p915) and extended data figure 2 (Created in BioRender. Tal, M. (2025) https://BioRender.com/m52s998) were created using BioRender. All other figures were created using Illustrator.

## FinnGen ethics statement

The FinnGen project is funded by two grants from Business Finland (HUS 4685/31/2016 and UH 4386/31/2016) and the following industry partners: AbbVie Inc., AstraZeneca UK Ltd, Biogen MA Inc., Bristol Myers Squibb (and Celgene Corporation & Celgene International II Sàrl), Genentech Inc., Merck Sharp & Dohme LCC, Pfizer Inc., GlaxoSmithKline Intellectual Property Development Ltd., Sanofi US Services Inc., Maze Therapeutics Inc., Janssen Biotech Inc, Novartis AG, and Boehringer Ingelheim International GmbH. Following biobanks are acknowledged for delivering biobank samples to FinnGen: Auria Biobank (www.auria.fi/biopankki), THL Biobank (www.thl.fi/biobank), Helsinki Biobank (www.helsinginbiopankki.fi), Biobank Borealis of Northern Finland (https://www.ppshp.fi/Tutkimus-ja-opetus/Biopankki/Pages/Biobank-Borealis-briefly-in-English.aspx), Finnish Clinical Biobank Tampere (www.tays.fi/en-US/Research_and_development/Finnish_Clinical_Biobank_Tampere), Biobank of Eastern Finland (www.ita-suomenbiopankki.fi/en), Central Finland Biobank (www.ksshp.fi/fi-FI/Potilaalle/Biopankki), Finnish Red Cross Blood Service Biobank (www.veripalvelu.fi/verenluovutus/biopankkitoiminta) and Terveystalo Biobank (www.terveystalo.com/fi/Yritystietoa/Terveystalo-Biopankki/Biopankki/). All Finnish Biobanks are members of BBMRI.fi infrastructure (www.bbmri.fi). Finnish Biobank Cooperative -FINBB (https://finbb.fi/) is the coordinator of BBMRI-ERIC operations in Finland. The Finnish biobank data can be accessed through the Fingenious® services (https://site.fingenious.fi/en/) managed by FINBB.

## All of Us Acknowledgements

We gratefully acknowledge All of Us participants for their contributions, without whom this research would not have been possible. We also thank the National Institutes of Health’s All of Us Research Program for making available the participant data [and/or samples and/or cohort] examined in this study. The All of Us Research Program is supported by the National Institutes of Health, Office of the Director: Regional Medical Centers: 1 OT2 OD026549; 1 OT2 OD026554; 1 OT2 OD026557; 1 OT2 OD026556; 1 OT2 OD026550; 1 OT2 OD 026552; 1 OT2 OD026553; 1 OT2 OD026548; 1 OT2 OD026551; 1 OT2 OD026555; IAA #: AOD 16037; Federally Qualified Health Centers: HHSN 263201600085U; Data and Research Center: 5 U2C OD023196; Biobank: 1 U24 OD023121; The Participant Center: U24 OD023176; Participant Technology Systems Center: 1 U24 OD023163; Communications and Engagement: 3 OT2 OD023205; 3 OT2 OD023206; and Community Partners: 1 OT2 OD025277; 3 OT2 OD025315; 1 OT2 OD025337; 1 OT2 OD025276. In addition, the All of Us Research Program would not be possible without the partnership of its participants.

## In Vivo ethics statement

*In vivo* studies performed at Stanford University followed the ethical care and use guidelines outlined in the Stanford University Administrative Panel on Laboratory Animal Care (APLAC) protocol 30109. All mice were housed in the Rodent Animal Facility at Stanford School of Medicine (Palo Alto, CA) which is accredited by the Association for Accreditation of Laboratory Animal Care (AAALAC).

*In vivo* studies performed at Massachusetts Institute of Technology were performed at the Department of Comparative (DCM) at the Massachusetts Institute of Technology (MIT) (Cambridge, MA). All procedures and care guidelines were approved by the MIT Committee on Animal Care (CAC) (Protocol#1221-087-24).

